# Coronavirus GenBrowser for monitoring the transmission and evolution of SARS-CoV-2

**DOI:** 10.1101/2020.12.23.20248612

**Authors:** Dalang Yu, Xiao Yang, Bixia Tang, Yi-Hsuan Pan, Jianing Yang, Guangya Duan, Junwei Zhu, Zi-Qian Hao, Hailong Mu, Long Dai, Wangjie Hu, Mochen Zhang, Ying Cui, Tong Jin, Cui-Ping Li, Lina Ma, Language translation team, Xiao Su, Guoqing Zhang, Wenming Zhao, Haipeng Li

**Author notes:** Joint Authors (Dalang Yu, Xiao Yang, Bixia Tang, Yi-Hsuan Pan, Jianing Yang, Guangya Duan).

## Abstract

Genomic epidemiology is important to study the COVID-19 pandemic and more than two million SARS-CoV-2 genomic sequences were deposited into public databases. However, the exponential increase of sequences invokes unprecedented bioinformatic challenges. Here, we present the Coronavirus GenBrowser (CGB) based on a highly efficient analysis framework and a movie maker strategy. In total, 1,002,739 high quality genomic sequences with the transmission-related metadata were analyzed and visualized. The size of the core data file is only 12.20 MB, efficient for clean data sharing. Quick visualization modules and rich interactive operations are provided to explore the annotated SARS-CoV-2 evolutionary tree. CGB binary nomenclature is proposed to name each internal lineage. The pre-analyzed data can be filtered out according to the user-defined criteria to explore the transmission of SARS-CoV-2. Different evolutionary analyses can also be easily performed, such as the detection of accelerated evolution and on-going positive selection. Moreover, the 75 genomic spots conserved in SARS-CoV-2 but non-conserved in other coronaviruses were identified, which may indicate the functional elements specifically important for SARS-CoV-2. The CGB not only enables users who have no programming skills to analyze millions of genomic sequences, but also offers a panoramic vision of the transmission and evolution of SARS-CoV-2.

## Introduction

Real-time tracking of the transmission and evolution of the severe acute respiratory syndrome coronavirus 2 (SARS-CoV-2) is essential for public health during the COVID-19 pandemic [1]. Since January 2020 more than two million genomic sequences have been deposited into public databases, such as National Center for Biotechnology Information (NCBI) GenBank [2], Global Initiative on Sharing All Influenza Data (GISAID) [3, 4]. The exponential increase of genomic sequences provides a great opportunity to monitor the transmission and evolution of SARS-CoV-2 but invokes unprecedented bioinformatic challenges.

Several web browsers have been developed to analyze the genomic data and track the COVID-19 pandemic. The UCSC SARS-CoV-2 Genome Browser was derived from the well-established UCSC genome-browser for visualization of nucleotide and protein sequences, sequence conservations, and many other properties of wild-type and variants of SARS-CoV-2 [5]. The WashU Virus Genome Browser provides Nextstrain-based phylogenetic-tree view and genomic-coordinate, track-based view of genomic features of viruses [6]. The pathogen genomics platform Nextstrain allows analysis of genomic sequences of approximately 4,000 strains of SARS-CoV-2 and investigation of its evolution [7], which cannot timely analyze millions of increasing genomic sequences. Therefore, new approaches are essential and indispensable to enable users easier to explore the large amount of SARS-CoV-2 genomic sequences.

In this study, we developed the Coronavirus GenBrowser (CGB). All the high-quality genomic sequences and the associated transmission-related metadata were timely analyzed to provide the latest panoramic view of the pandemic. To investigate a local transmission, the data can be easily filtered according to countries, regions, keywords, and the collection date of viral strains. Thus, even if users have no any programming skills, the CGB enables them to efficiently explore millions of SARS-CoV-2 genomic sequences and monitor the global/local transmission and evolution of SARS-CoV-2. All the pre-analyzed genomic mutations and the associated metadata can be easily downloaded, re-analyzed and re-shared. Since a cleaned genome alignment with almost no ambiguous nucleotide sites can be easily re-constructed from the CGB core data file, the CGB may provide a great convenience for the society to study viral evolution further.

## Material and methods

Brief descriptions of material and methods are included in this section. Detailed descriptions are provided as Supplemental Materials.

### Data quality control and distributed genome alignments

SARS-CoV-2 genomic variations were obtained from the 2019nCoVR database [8] established by China National Center for Bioinformation (CNCB) [9], as an integrated resource based on Global Initiative on Sharing All Influenza Data (GISAID) [3, 4], National Center for Biotechnology Information (NCBI) GenBank [2], China National GeneBank DataBase (CNGBdb) [10], the Genome Warehouse (GWH) [11], and the National Microbiology Data Center (NMDC, https://nmdc.cn/). Detailed information on this database is available at https://bigd.big.ac.cn/ncov/release_genome. All SARS-CoV-2 strains were isolated from humans, and quality control was applied to obtain high-quality SARS-CoV-2 genomic sequences (Figure S1, S2). Because of the explosion in SARS-CoV-2 genomic data, the distributed alignment system was developed to enable daily update (Figure 1), which reduces the total alignment time complexity to 𝒪(*n*), where 𝒪(.) is a linear function, and *n* is the number of viral strains.

**Figure 1.**
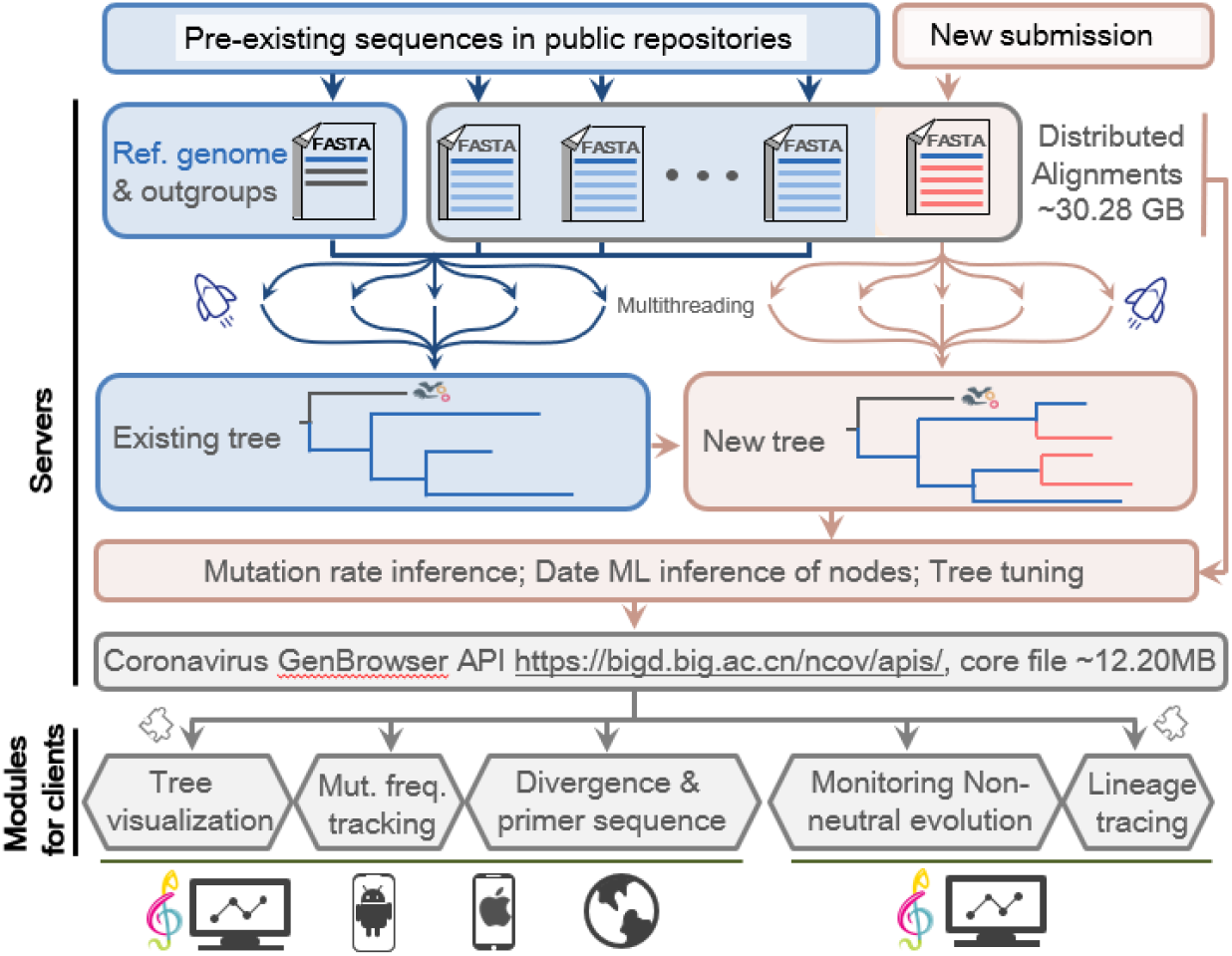
Timely updates of SARS-CoV-2 genomic data and visualization framework of Coronavirus GenBrowser. The core file includes the pre-analyzed genomic mutations of SARS-CoV-2 and the associated metadata. All timely-updated data can be freely accessed at https://bigd.big.ac.cn/ncov/apis/.

### Reconstruction and timely update of the annotated phylogenetic tree

Before October 2020 all high-quality sequences in distributed alignments were analyzed as a whole and used to reconstruct the evolutionary tree. After October 2020 the efficiency of this process became too low to perform timely updates. Therefore, new trees were reconstructed by appending new sequences to existing tree. Ambiguous and missing nucleotides were imputed by incorporation of the neighboring lineages (Figure S3) and mutations in strains of each branch are recapitulated according to the principle of parsimony [12, 13]. A highly effective maximum-likelihood method (TreeTime) is used to determine the dates of internal nodes [14] as it allows fast inference by “the post- and pre-order traversals” with tabulated key values for back tracing. This algorithm was implemented in the CGB with very minor revisions (Figures S4, and S5). The genome-wide mutation rate was also timely calculated.

### Displaying SARS-CoV-2 genomic mutations in tree-based format

Similar to NextStrain [7] and the WashU Virus Genome Browse [6], the CGB uses a tree-based file format to store SARS-CoV-2 genomic mutations. The head of the core data file contains the data version, the updated date, the genomic region analyzed, and the mutation rate estimated for each gene. The core file also contains information on collection date, gender and age of patient, location for each strain, mutations, and inferred date for each internal node. To allow fast access to the data, redundant information has been minimized.

### Visualization of the huge evolutionary tree by movie-maker strategy

When visualizing the huge evolutionary tree, the most of lineages are invisible because lineages overlap each other. Thus, a movie-maker strategy was implemented to efficiently visualize huge evolutionary tree, in which only surface lines will be painted. If the tree is zoomed in, only the visible sub-area of the tree will be painted. Using this strategy, millions of lineages can be visualized effectively (Figure S6).

### CGB binary nomenclature and data searching

A number of different naming systems have been proposed [15, 16], but these systems only name a few internal nodes or branches. As there are a large number of internal nodes on the huge evolutionary tree, the CGB binary nomenclature was developed following the most recent common ancestor (MRCA) concept (Figure S11) to obtain CGB ID for each node. CGB ID can be used to search a specific lineage. Isolate names, accession numbers, and mutations are also searchable.

### Mutation analysis

A root-to-tip linear regression method [17] was used to estimate the mutation rate of SARS-CoV-2. For each strain with a different collection date in a tip-dated time tree, the number of mutations, including that of recurrent mutations, was counted subsequent to the appearance of MRCA. To avoid the effect of recombination, recombination flag is labeled for each mutation by analyzing hybrid genomic structure (Figure S12).

### Lineage tracing

For lineage tracing, genomic sequences of SARS-CoV-2 strains collected from patients or environments are used as the queries. These query sequences should be aligned with the reference genomic sequence of SARS-CoV-2 (GenBank accession number: NC_045512) [18] A very fast algorithm was implemented to count the difference between a query sequence and the genomic sequence of a node. For one query, nodes with the least difference are considered as its candidate targets.

### Detection of branch-specific accelerated evolution of SARS-CoV-2

To detect branch-specific accelerated evolution, each internal branch of the SARS-CoV-2 tree was examined. For each internal branch, the observed number of mutations of the *i*-th gene (*γ*_*obs,i*_) was compared with the expected number of mutations of the same gene (*γ*_*exp,i*_). The significance level of acceleration was determined by Poisson probability [19, 20]. It is a one-tailed test. The condition *t* > 10 (days) was used for detection of branch-specific accelerated evolution of SARS-CoV-2.

### Detection of on-going selection of SARS-CoV-2

To detect on-going positive selection, allele frequency trajectory with an S-shaped curve was examined (Table S3, Figure S13, S14). To reduce the impact of hitchhiking by neutral mutation, only non-synonymous mutations were analyzed, although non-coding mutations [21] can also be beneficial.

### Local analysis for new genomic data of SARS-CoV-2

The public global genomic data can be freely downloaded and analyzed together with new genomic data before these new sequences are integrated into the CGB by the CGB team members. This function ensures that a timely analysis can be easily performed.

### Data source

For genomic sequence alignments, high quality SARS-CoV-2 genomic variations were obtained from the 2019nCoVR database [8, 22], which is an integrated resource based on GenBank, GISAID [3, 4], China National GeneBank DataBase (CNGBdb) [10], the Genome Warehouse (GWH) [11], and the National Microbiology Data Center (NMDC, https://nmdc.cn/).

## Results

### The construction of a million level evolutionary tree

After quality control, 1,002,739 high-quality genomic sequences were obtained for subsequent analyses. The number of identified high- and low-quality genomes in each month is shown (Figure S2). To allow timely analysis of a large number of sequences, we first solved the problem that all viral genomic sequences have to be re-aligned when nucleotide sequences of new genomes become available. This is extremely time-consuming. With the distributed alignment system (Figure 1), we dramatically reduced the total time required for the alignment. We also built the evolutionary tree on the existing tree with new genomic data in order to reduce the complexity of tree construction. With these modifications, a tremendous evolutionary tree can be reconstructed for each update, millions of SARS-CoV-2 genomic sequences can be timely analyzed, and data can be easily shared, reanalyzed and re-constructed (Figure 1).

For the huge evolutionary tree, mutations on each branch were identified according to the principle of parsimony [12, 13] and the dates of internal nodes were inferred with very minor revisions of a highly effective maximum-likelihood method (TreeTime) [14]. The pre-analyzed genomic mutations of SARS-CoV-2 and the associated metadata are shared to the general public in a tree-based CGB format. The size of distributed alignments is 30.28 GB for the 1,002,739 SARS-CoV-2 genomic sequences. The tree-based data format allows the compression ratio to reach 2,541:1, meaning that the size of compressed core file containing the pre-analyzed genomic mutations and associated metadata is as small as 12.20 MB with zip compression (Figure 1). Whenever necessary, cleaned genomic sequences with almost no ambiguous nucleotide sites can be reconstructed for viral isolates. Thus, this approach ensures low-latency access to the data and enables fast sharing and re-analysis of a large number of SARS-CoV-2 genomic variants.

### Highly efficient visualization of the tree and tracks

To efficiently visualize the results, a movie-maker strategy was implemented for painting the evolutionary tree, only elements shown on the screen and visible to the user are painted. This design makes the visualization process highly efficient, and the evolutionary tree of more than one million strains can be visualized. It takes about one second for the visualization process in different operation systems (Table S1).

### The convenience of tree operation

The CGB is also a highly efficient platform to search or filter variants based on transmission-related metadata (Figure 2). Useful interactive functionalities were developed to navigate users through the huge tip-dated evolutionary tree. First, users can easily search internal branches or variants with certain mutations, or isolate names of virus. There are 400,298 internal branches in the evolutionary tree (*n* = 1,002,739), and each branch has been named by CGB binary nomenclature (*i*.*e*., CGB ID) (Figure S11) and is searchable. Thus, different Variants of Concerns (VOCs) can be easily identified and visualized on the huge evolutionary tree (Table 1). Second, users can easily filter out the data according to the collection date, the country/region, and the gender and age groups. Third, the visualization of a sub-clade in another tab is allowed. Forth, different annotations are provided to mark the clades of interest. At last, coordinated annotation tracks are provided to show genome structure and key domains, allele frequencies, sequence similarity between various coronavirus, multi-genome alignment and primer sets for detection of SARS-CoV-2 (Figure S7-10). All those features have brought great convenience for users.

**Figure 2.**
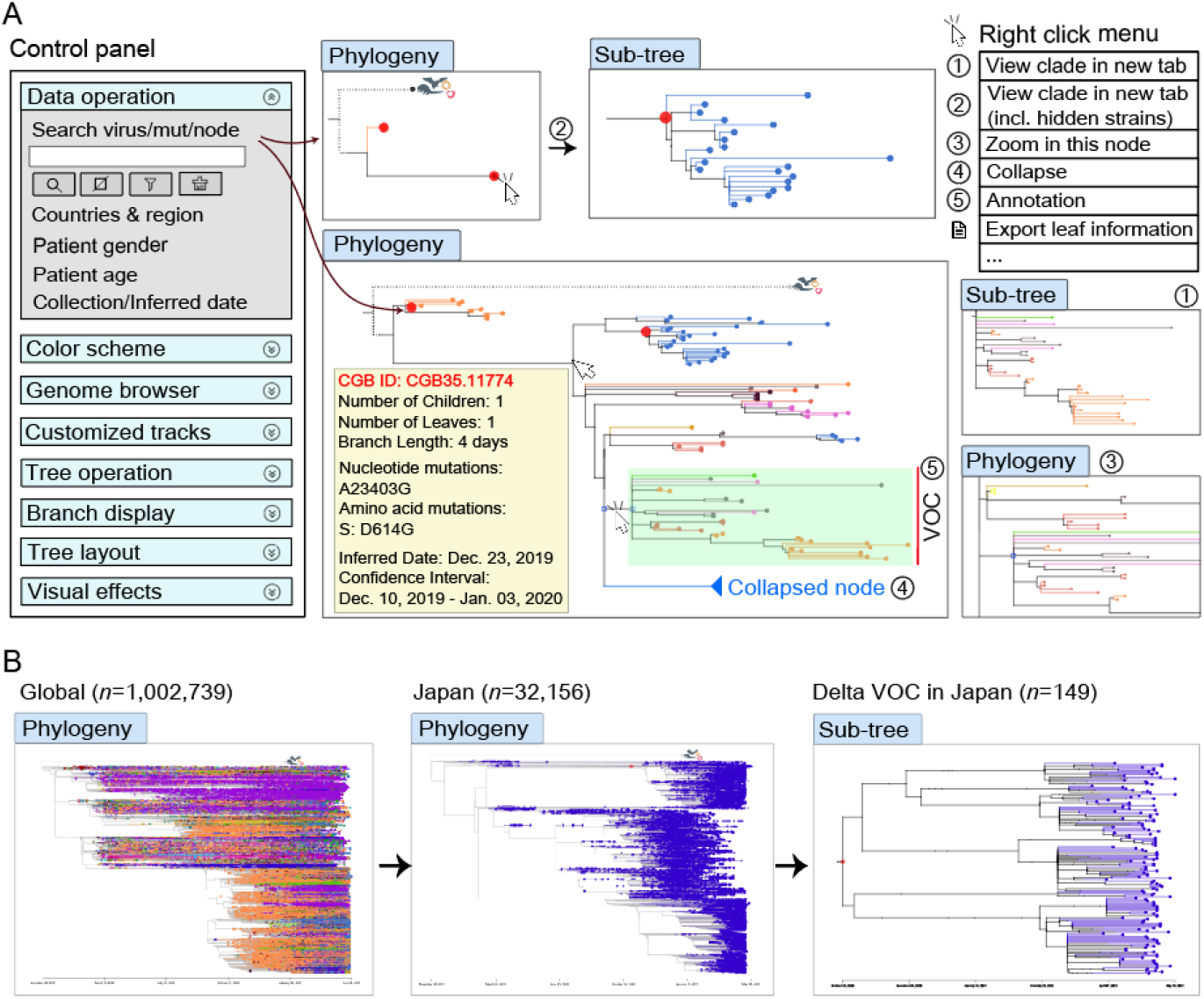
An illustration of CGB interactive functionalities. (A) A sketch of the tree visualization panel to present the major CGB functions. Strains, mutations, and CGB IDs can be searched. The searched nodes are highlighted and can be visualized individually. A number of frequently-used menus are presented at the top right corner. (B) An example for how to view the transmission and evolution of the Delta VOC in Japan. First, users can filter for strains collected from Japan. Search the CGB ID of the Delta VOC (CGB531065.580055) and then right click the highlighted node and choose the menu item of View clade in new tab in the pop-up menu.

**Table 1.**
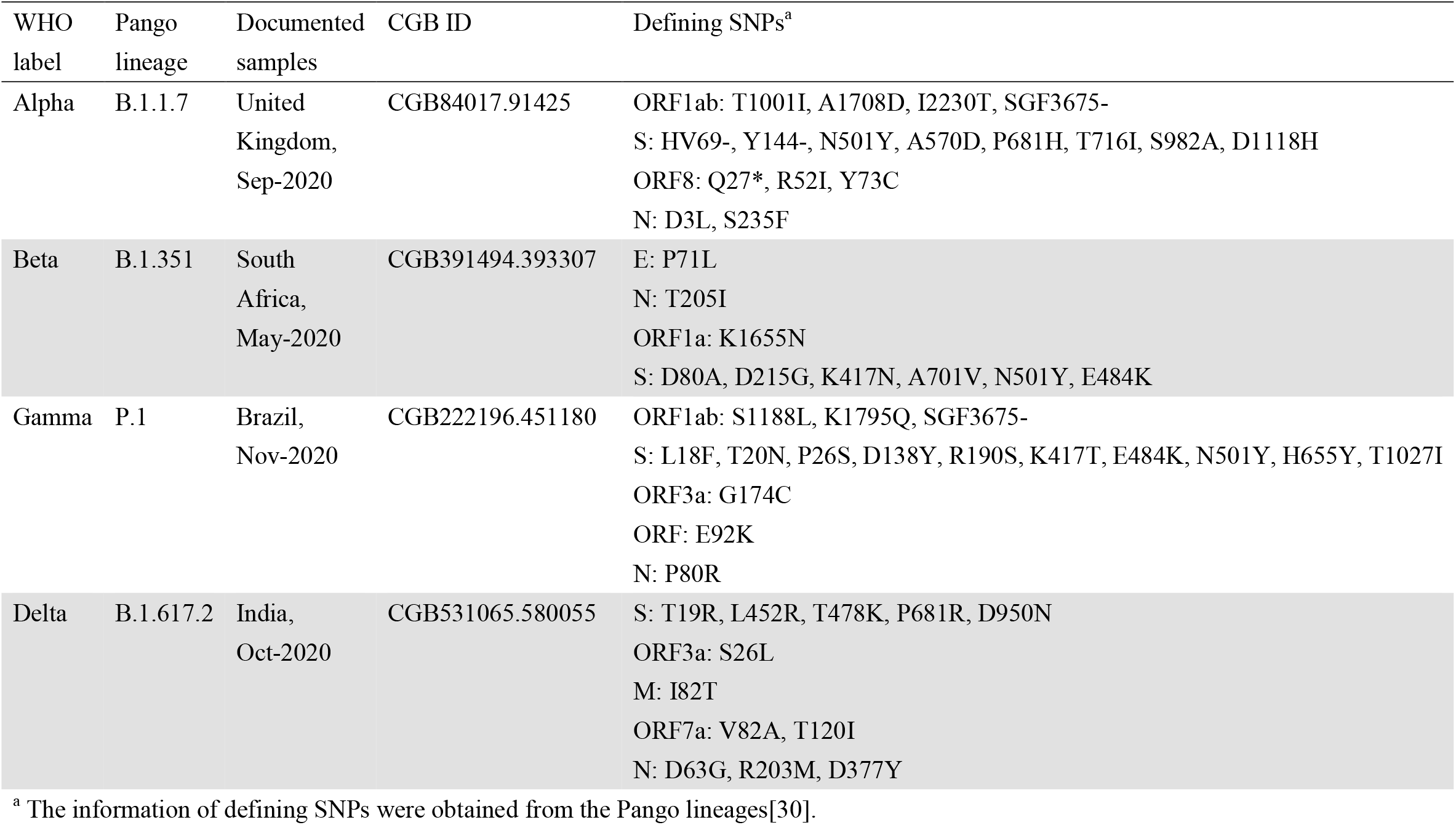
CGB ID for Variant of Concern (VOC).

### Transmission case studies through CGB

Based on the large data volume and user-friendly CGB, many analyses can be quickly conducted. Take the spread of VOCs in India as an example. Among the 1,002,739 strains, there were 3,349 ones sampled in India. Nearly all major SARS-CoV-2 lineages can be found in India in different stages of the pandemic (Figure 3). By searching the CGB IDs of the VOCs (Table 1), the clades of VOCs were identified. In total, there were 464 (464/3,349=13.85%) Delta strains, 185 (185/3,349=5.52%) Alpha strains, and 11 (11/3,349=0.32%) Beta strains in the Indian sample. The ratios change when only considering recent viral strains collected after Apr 01, 2021. There were 589 India samples after date filtering. Among them, there were 364 (364/589=61.80%) Delta strains, 55 (55/589=9.34%) Alpha (B.1.1.7) strains and one (1/589=0.17%) Beta strains. Thus, the Delta variant increased more rapidly than others, and the most recent infections in India are caused by the Delta (B.1.617.2) VOC.

**Figure 3.**
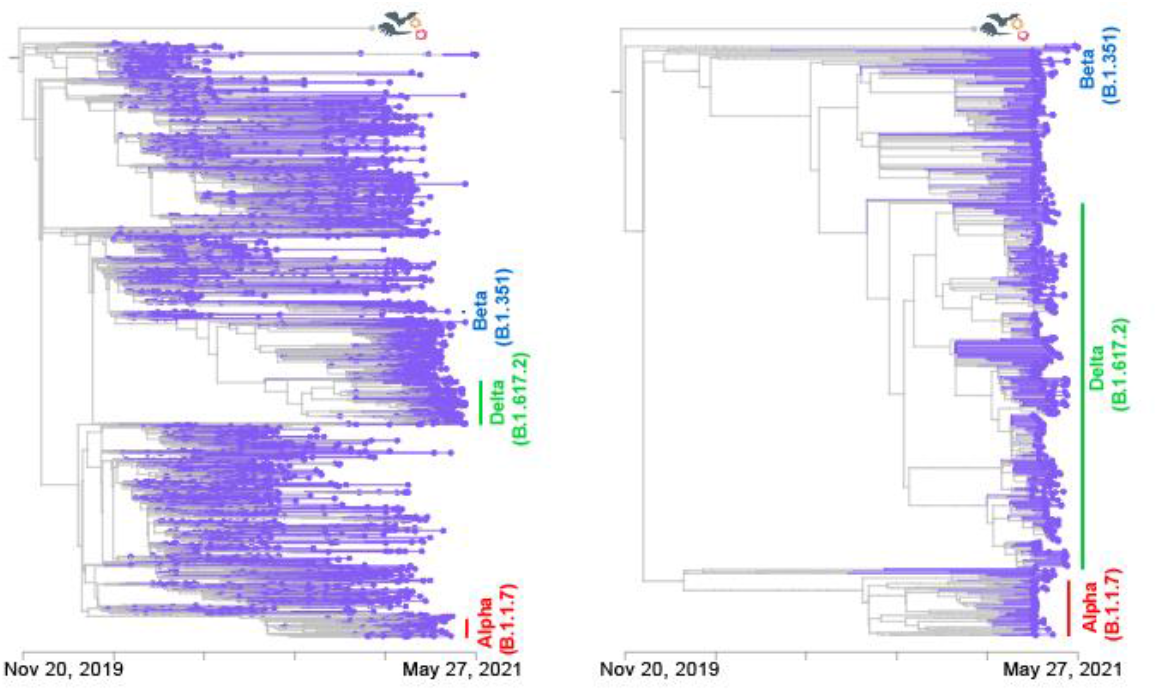
Proportion change of VOCs in India. The tree on the left shows all India samples of 1,002,739 SARS-CoV-2 strains and the tree on the right shows India samples after Apr 01, 2021. Different VOC are annotated in different colors, Alpha (B.1.1.7) VOC in red, Delta (B.1.617.2) VOC in green and Beta (B.1.351) VOC in blue, no Gamma are found in India.

Another three examples were provided to show that the CGB is an efficient platform to investigate local and global transmission of COVID-19 (Figure 4). To trace the origin of a local COVID-19 outbreak, the lineage tracing was implemented in the CGB. The closest nodes were revealed for the three outbreaks in China during this year which indicates different origin of the three outbreaks. Their neighboring strains can be viewed individually and further investigated. The analysis is extremely fast and can be performed on a desktop computer. Therefore, the CGB is a highly efficient platform to investigate the origin of a local COVID-19 outbreak.

**Figure 4.**
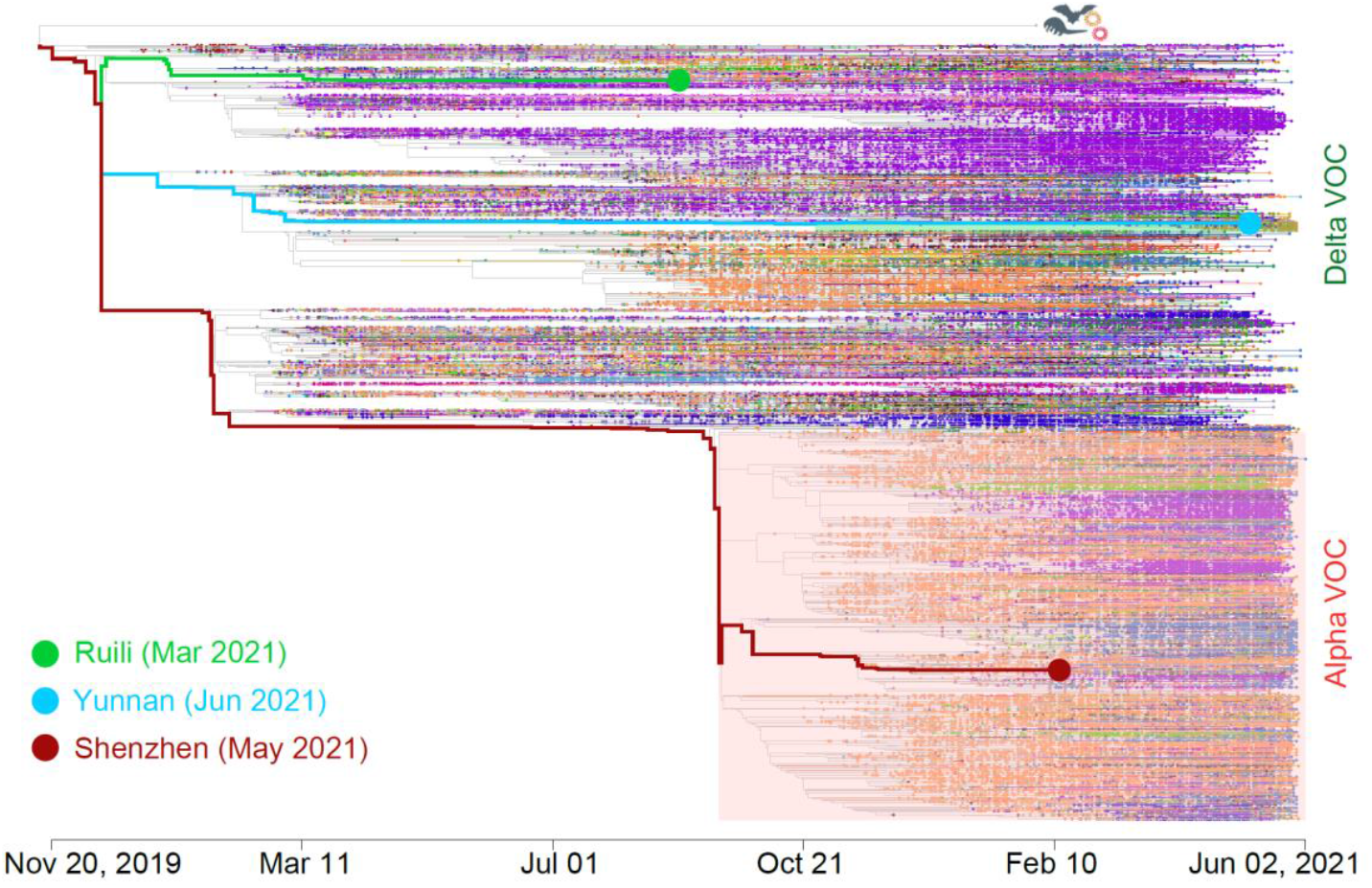
Tracing the origin of three local outbreak of COVID-19 in China. One sequence sampled during each outbreak is used as the query sequence which is not included in the CGB dataset version used in analysis. Their closest targets were marked with colored dot and the evolutionary paths were highlighted. The GISAID IDs for the queries are EPI_ISL_1595852 (Ruili), EPI_ISL_2834004 (Yunnan), and EPI_ISL_2405168 (Shenzhen).

### Mutation analysis

The CGB also estimates the mutation rate of whole genome and each gene (Table S2). Applying 1,002,739 genomic sequences, the estimated genome-wide mutation rate is 1.0794 × 10^−3^ per nucleotide per year. The mutation rate is variable for different genes (Table S2).

We also found that the mutation rate could be different among sites. Using the CGB core data file, we conducted a 10-base sliding window analysis with a sliding step of one base and identified fine-scaled mutation cold spots along the viral genome, indicating the genomic regions with mutation rate significantly lower than the average mutation rate of the entire genome. In total, 657,074 (recurrent) mutations were identified and 868 mutation cold spots were found with a false discovery rate (FDR) corrected *P*-value < 0.01 (Figure 5, Supplemental excel file). The coldest spot is located in ORF1ab, which encodes nsp13 helicase (nucleotides 16,294 – 16,307) (FDR corrected *P*-value = 4.79 × 10^−46^). Interestingly, it has been found that sequence conservation is restricted to ORF1ab:nsp10-13 among 14 coronaviruses [23, 24]. It indicates that nsp13 helicase might be essential for coronaviruses and SARS-CoV-2. Moreover, among the 868 mutation cold spots, there are 75 conserved spots in SARS-CoV-2, but not conserved among other coronaviruses. These SARS-CoV-2 specific conserved elements may play key roles for SARS-CoV-2 specific functions.

**Figure 5.**
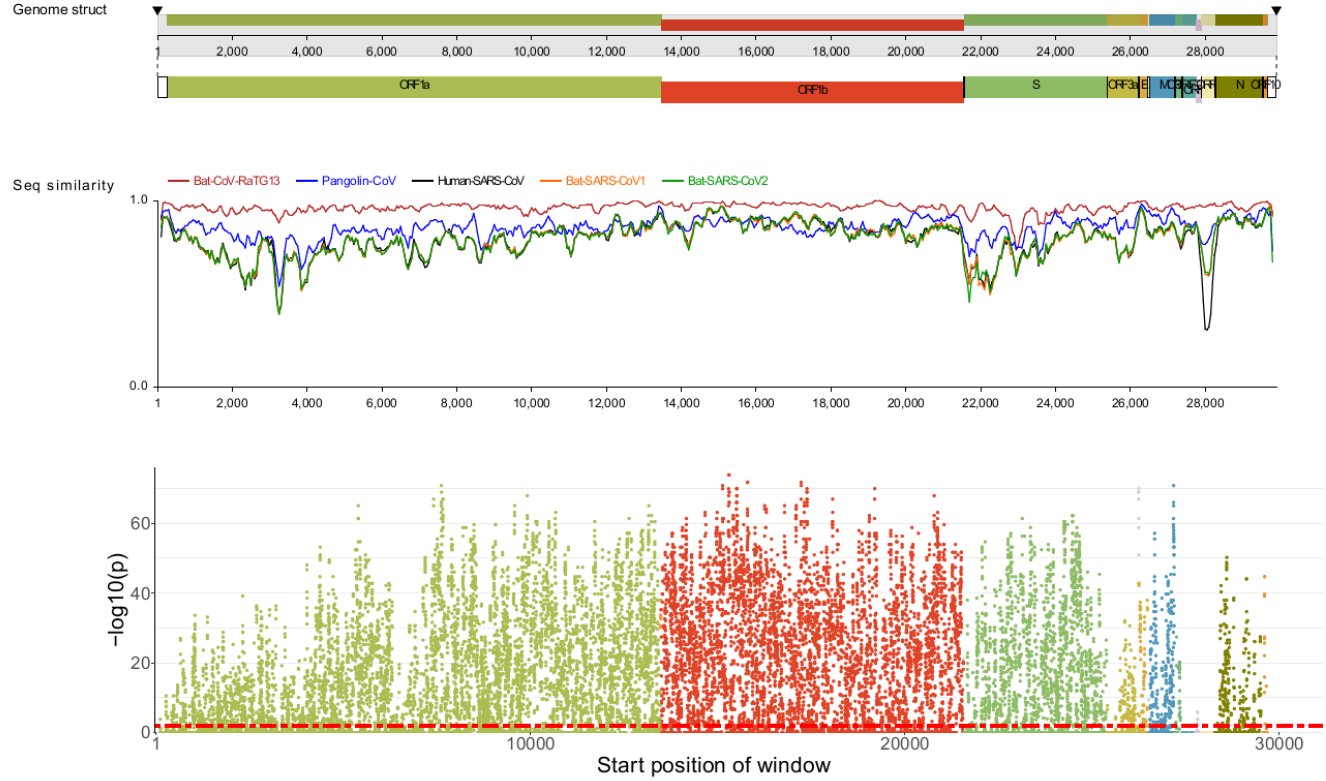
Manhattan plot of mutation cold spots in the genome of SARS-CoV-2. Results of genome-wide scan for mutation cold spots are shown in Manhattan plot of significance against SARS-CoV-2 reference genomic locations. In total, 1,002,739 high quality genomic sequences were analyzed. Each dot represents one window. *P*-values are FDR-corrected. The dotted red line denotes FDR-corrected *P*-value < 0.01. Dots above the line represent mutation cold spots. Genomic structure and sequence similarity between SARS-CoV-2 reference genome (NC_045512.2)[18] and the genomes of five other coronaviruses are shown above the Manhattan plot.

### The detection of accelerated evolution

The CGB provides a module to detect branch-specific accelerated evolution of SARS-CoV-2. We found that within 186,746 internal branches with *t* > 10 (days) on the evolutionary tree (*n* = 1,002,739), 70 branches were detected to have a genome-wide accelerated evolution (FDR corrected *P* < 0.05), 332 branches were detected to have an accelerated evolution of ORF1ab (FDR corrected *P* < 0.05), and two branches was found to have an accelerated evolution of the spike gene (FDR corrected *P* < 0.05) (Supplemental excel file). These evolution-accelerated variants can be used for future studies.

### The detection of on-going positive selection

The mutation frequency trajectory for each mutation can be easily visualized by using the CGB. It has a module to detect on-going positive selection based on S-shaped frequency trajectory of a selected allele (Figures S13, S14). It has been shown that the SARS-CoV-2 variant with G614 spike protein has a fitness advantage [25]. Our analysis using the CGB confirmed this finding even when the G614 frequency was very low (< 10%) (Figure S15), indicating that the CGB can detect putative advantageous variants before they become widely spread. The CGB also predicted an increase in the frequency of S:p.P681R of the Delta VOC (Figure S15), suggesting that variants with the mutation may be advantageous. S:p.P681R is located on the spike S1/S2 cleavage site, and another mutation (S:p.P681H) on the same position has been found to be advantageous [26] and may interact with other mutations [21] in the Alpha VOC. Based on 1,002,739 samples, the CGB detected 13 putative advantageous mutations in the spike protein (Table S4). However, as an increase in mutation frequency could be due to sampling bias and epidemiological factors [25], putative advantageous variants should be closely monitored.

## Conclusion

In this study, we developed an effective surveillance tool for the transmission and evolution of SARS-CoV-2. A highly efficient visualization module is established, and rich interactive operations are allowed to explore the annotated evolutionary tree. We investigated four local COVID-19 outbreaks by the searching and lineage tracing functionalities. We also implemented a new method to detect on-going positive selection for each viral nonsynonymous mutation. The branches with accelerated or reduced evolutionary rate are identified to provide a real time tracking on the change of evolutionary rate, which could reveal epidemic factors affecting the viral transmission.

The CGB provides an efficient way for clean data sharing. It could be difficult for researchers to download all the raw genomic sequences, perform data quality control and analyze the large amount of data by their own. By examining literatures related to this topic [27, 28], most of those studies have similar methods to prepare alignments, build phylogenetic tree, infer mutations and date each internal node. The data preparation procedure is tedious but may require some skills and is often time consuming when the sample size is extremely large. Nevertheless, by downloading the pre-analyzed CGB data files, users can have all those clean data in a few minutes. To promote a timely analysis of newly sequenced genomic data, users can perform a local analysis to analyze these new data, together with the public genomic data globally sampled. Thus, the CGB provides a convenient way to study the evolution of SARS-CoV-2 and monitor its transmission.

During the analysis, we noticed that a very small percentage of sequences have abnormal collection dates which could severely skew the evolutionary tree. After examining all possible reasons, it is likely due to that the year of collection date was incorrectly filled in the most cases. We then deleted these sequences although their sequence quality is high. Thus, we would suggest that researchers could pay attention on the year of collection date when submitting their sequences.

The public science education is extremely important for the anti-epidemic to show that SARS-CoV-2 has been evolving. Therefore, a web-based CGB was also developed. It is a simplified version of CGB that provides a convenient way to access the data via a web browser, such as Google Chrome, Firefox, and Safari (Figure S6). For educational purpose, nine language versions (Chinese, English, German, Japanese, French, Italian, Portuguese, Russian, and Spanish) are available. The web-based CGB package can be downloaded and reinstalled on any websites. Two pre-installed websites are provided (https://www.biosino.org/genbrowser/ and https://ngdc.cncb.ac.cn/genbrowser/). For the scientific community, it is highly recommended to download the eGPS software to use the CGB (http://www.egps-software.net/egpscloud/eGPS_Desktop.html). Overall, the CGB is frequently updated which provides a timely panoramic vision of the global and local transmission and evolution of SARS-CoV-2.

## Data Availability

All timely-updated data are freely available at https://bigd.big.ac.cn/ncov/apis/. The desktop standalone version provides the full function of CGB and has a plug-in module for the eGPS software (http://www.egps-software.net/egpscloud/eGPS_Desktop.html).

## Funding

This work was supported by a grant from the National Key Research and Development Project [No. 2020YFC0847000]. Funding for open access charge: National Key Research and Development.

## Acknowledgements

We thank Ya-Ping Zhang for providing valuable advice and encouragement and the researchers who generated and deposited sequence data of SARS-CoV-2 in GISAID, GenBank, CNGBdb, GWH, and NMDC making this study possible.

## Data availability

All timely-updated data are freely available at https://bigd.big.ac.cn/ncov/apis/. The desktop standalone version (Figure S6A) provides the full function of CGB and has a plug-in module for the eGPS software (http://www.egps-software.net/egpscloud/eGPS_Desktop.html) [29].

## Members of the language translation team

German: Ning He^6^, Jing Lv^6^, Ting Peng^6^

Italian: Ting Zhou^6^, Nan Yang^6^, Siyi Hou^6^

Portuguese: Huang Li^6^, Jingxuan Yan^6^, Chenglin Zhu^6^, Wenjing Liu^6^

Russian: Yuhong Guan^6^, Huanxiao Song^6^

Spanish: Qin Zhou^6^, Han Gao^6^, Jinglan He^6^, Tiantian Li^6^, Ruiwen Fei^6^, Shumei Zhang^6^

French: Yuyuan Guo^6^

## Conflict of interest

The authors declare no competing interests.

## Notes

### Competing Interest Statement

The authors have declared no competing interest.

### Summary of Updates

The manuscript was re-written to focus on the Coronavirus GenBrowser (CGB) itself. The readers may feel more easy to understand how and why the CGB was developed.

## References

1. Fineberg HV, Wilson ME. Epidemic science in real time, Science 2009;324:987.

2. Sayers EW, Beck J, Bolton EE et al. Database resources of the National Center for Biotechnology Information, Nucleic Acids Res 2021;49:D10–D17.

3. Shu YL, McCauley J. GISAID: Global initiative on sharing all influenza data - from vision to reality, Eurosurveillance 2017;22:2–4.

4. Elbe S, Buckland-Merrett G. Data, disease and diplomacy: GISAID’s innovative contribution to global health, Glob Chall 2017;1:33–46.

5. Fernandes JD, Hinrichs AS, Clawson H et al. The UCSC SARS-CoV-2 Genome Browser, Nat Genet 2020;52:986–991.

6. Flynn JA, Purushotham D, Choudhary MNK et al. Exploring the coronavirus pandemic with the WashU Virus Genome Browser, Nat Genet 2020;52:986–1001.

7. Hadfield J, Megill C, Bell SM et al. Nextstrain: real-time tracking of pathogen evolution, Bioinformatics 2018;34:4121–4123.

8. Zhao W-M, Song S-H, Chen M-L et al. The 2019 novel coronavirus resource, Hereditas (Beijing) 2020;42:212–221.

9. Xue YB, Bao YM, Zhang Z et al. Database resources of the National Genomics Data Center, China National Center for Bioinformation in 2021, Nucleic Acids Res 2021;49:D18–D28.

10. Chen F, You L, Yang F et al. CNGBdb: China National GeneBank DataBase, Hereditas (Beijing) 2020;42:799–809.

11. Chen M, Ma Y, Wu S et al. Genome Warehouse: A public repository housing genome-scale data, Genomics Proteomics Bioinformatics 2021.

12. Sankoff D. Minimal mutation trees of sequences., SIAM J Appl Math 1975;28:35–42.

13. Hartigan JA. Minimum mutation fits to a given tree, Biometrics 1973;29:53–65.

14. Sagulenko P, Puller V, Neher RA. TreeTime: Maximum-likelihood phylodynamic analysis, Virus Evol 2018;4:vex042.

15. Tang X, Wu C, Li X et al. On the origin and continuing evolution of SARS-CoV-2, Natl Sci Rev 2020;7:1012–1023.

16. Forster P, Forster L, Renfrew C et al. Phylogenetic network analysis of SARS-CoV-2 genomes, Proc Natl Acad Sci USA 2020;117:9241–9243.

17. Bouckaert R, Vaughan TG, Barido-Sottani J et al. BEAST 2.5: An advanced software platform for Bayesian evolutionary analysis, PLoS Comput Biol 2019;15.

18. Wu F, Zhao S, Yu B et al. A new coronavirus associated with human respiratory disease in China, Nature 2020;579:265–269.

19. Ohta T, Kimura M. On the constancy of the evolutionary rate in cistrons, J Mol Evol 1971;1:18–25.

20. Wang Y, Dai G, Gu Z et al. Accelerated evolution of an Lhx2 xenhancer shapes mammalian social hierarchies, Cell Res 2020;30:408–420.

21. Yang J, Zhang G, Yu D et al. A Kozak-related non-coding deletion effectively increases B.1.1.7 transmissibility, bioRxiv 2021.

22. Gong Z, Zhu J-W, Li C-P et al. An online coronavirus analysis platform from the National Genomics Data Center, Zool Res 2020;41:705–708.

23. Ruan YJ, Wei CL, Ee LA et al. Comparative full-length genome sequence analysis of 14 SARS coronavirus isolates and common mutations associated with putative origins of infection, Lancet 2003;361:1779–1785.

24. He JF, Peng GW, Min J et al. Molecular evolution of the SARS coronavirus during the course of the SARS epidemic in China, Science 2004;303:1666–1669.

25. Korber B, Fischer WM, Gnanakaran S et al. Tracking changes in SARS-CoV-2 Spike: Evidence that D614G increases infectivity of the COVID-19 virus, Cell 2020;182:812–827.

26. Rambaut A, Loman N, Pybus O et al. Preliminary genomic characterisation of an emergent SARS-CoV-2 lineage in the UK defined by a novel set of spike mutations, virological.org 2020:https://virological.org/t/preliminary-genomic-characterisation-of-an-emergent-sars-cov-2-lineage-in-the-uk-defined-by-a-novel-set-of-spike-mutations/563.

27. Hodcroft EB, Zuber M, Nadeau S et al. Spread of a SARS-CoV-2 variant through Europe in the summer of 2020, Nature 2021.

28. Deng XD, Gu W, Federman S et al. Genomic surveillance reveals multiple introductions of SARS-CoV-2 into Northern California, Science 2020;369:582–587.

29. Yu D, Dong L, Yan F et al. eGPS 1.0: comprehensive software for multi-omic and evolutionary analyses, Natl Sci Rev 2019;6:867–869.

30. O’Toole Á, Hill V, Pybus OG et al. Tracking the international spread of SARS-CoV-2 lineages B.1.1.7 and B.1.351/501Y-V2, Wellcome Open Res 2021;6:121.

