## Supplemental methods and materials for "Coronavirus GenBrowser for monitoring the transmission and evolution of SARS-CoV-2"

†Joint Authors.

### Data quality control

SARS-CoV-2 genomic sequences were obtained from the 2019nCoVR database[1] established by China National Center for Bioinformation (CNCB). Detailed information on this database is available at https://bigd.big.ac.cn/ncov/release_genome. All SARS-CoV-2 isolates are from humans. To obtain high-quality SARS-CoV-2 genomic sequences, quality control measures were applied (Figure S1).

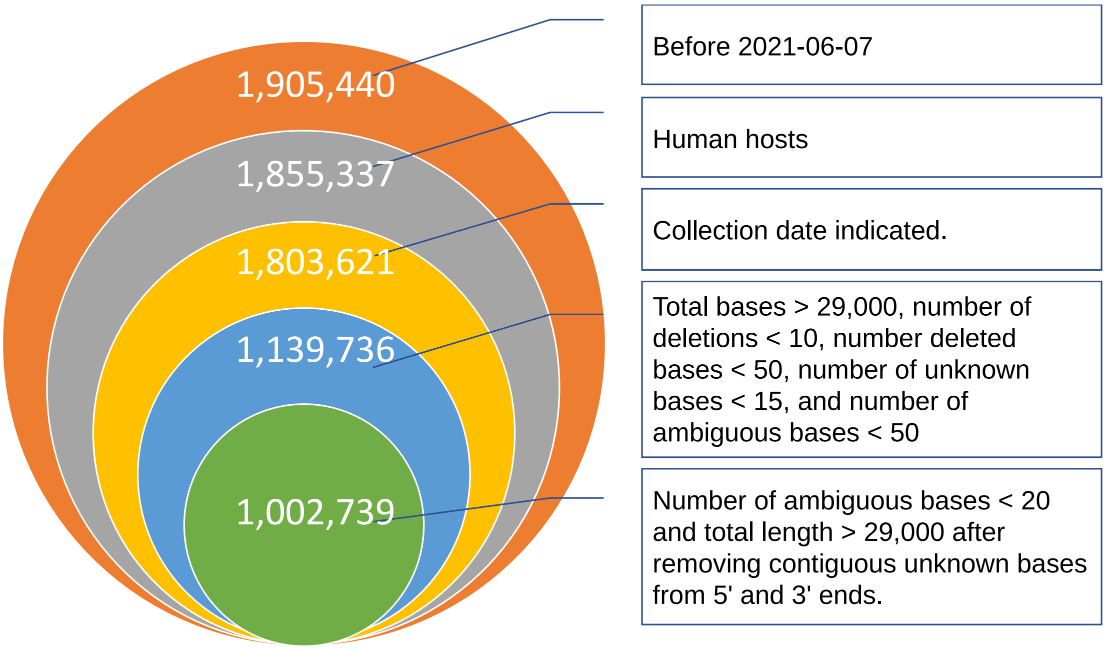

**Figure S1. Quality control pipeline.** The value in each circle is number of sequences identified in the quality control performed on 7 June, 2021.

The following criteria were used to select high quality sequences. First, the collection date of each strain is indicated. Second, the sequence length is longer than 29,000 bases, and the genome contains all protein-coding genes. Third, a gap found by sequence alignment is considered as one deletion, the number of deletions is <10, and the number of deleted bases is < 50. Forth, the number of unknown bases (Ns) is < 15, and the number of ambiguous bases (Ds) is < 50. Fifth, the length of the genome is longer than 29,000 bases after removing contiguous unknown bases from 5’ and 3’ ends. Sixth, as analysis of 23,336 genomes revealed that 5% of the genomes contain more than 19 ambiguous (Ds) and unknown (Ns) bases, a high-quality sequence must have a total number of ambiguous and unknown bases < 20.

After applying these criteria, 1,002,739 high-quality genomic sequences were identified and used for subsequent analyses, unless noted otherwise. The number of identified high- and low-quality genomes in each month is shown in Figure S2.

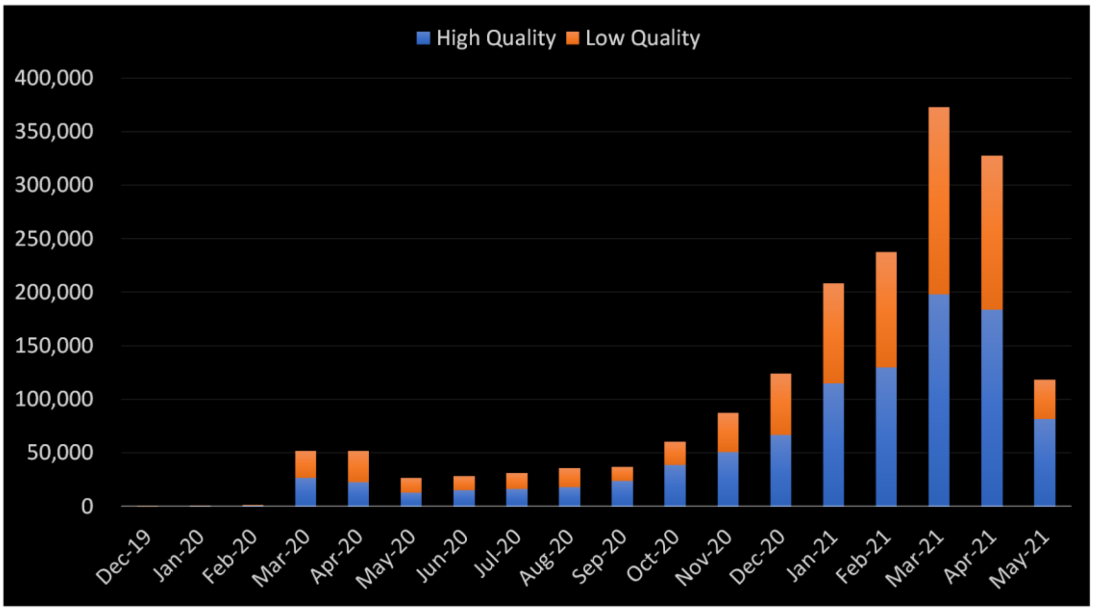

**Figure S2. Number of high- and low-quality SARS-CoV-2 genomic sequences at various time points.**

### Distributed genome alignments

Genome alignment was performed using the software MAFFT[2] with parameters "--auto --addfragments" after dividing input sequences into reference (GenBank accession number: NC_045512)[3] and others. Because of the explosion in SARS-CoV-2 genomic data, it is nearly impossible to perform daily update with the currently available analysis framework. To solve this problem, the distributed alignment system was developed (Figure 1), which reduces the total alignment time complexity to $\mathcal{O}(n)$, where $\mathcal{O}(.)$ is a linear function, and $n$ is number of viral strains. In this study, each alignment contained approximately 5,000 genomic sequences, including the reference SARS-CoV-2 sequence (NC_045512)[3]. To generate the outgroup alignment file, the reference sequence (NC_045512)[3] was aligned with the sequences of two outgroups: bat coronavirus RaTG13[4] and pangolin coronavirus PCoV-GX-P1E[5].

### Ancestral alleles of SARS-CoV-2

In total, 272 SARS-CoV-2 strains were collected before 31 January, 2020. These strains were collectively named “early samples” in this study. To detect ancestral alleles, the region between nucleotide positions 100 and 29,800 of each genome was examined. Compared to the reference sequence (NC_045512)[3], 28,846 monomorphic and 855 polymorphic sites were detected in the genomes of early samples, and the ancestral alleles for those sites are determined. Upon further comparison with the sequences of the two outgroups (RaTG13 and PCoV-GX-P1E)[4, 5], the majority of major alleles in 827 (96.7%) of the 855 polymorphic sites were found to be identical to the alleles in the outgroup genomes. Among the 28 unique polymorphic sites, minor alleles in 26 sites were found to be rare with a frequency less than 0.06, suggesting that the major alleles in these 26 sites in the early samples are ancestral. The frequencies of two major alleles *8,782C* and *28,144T* are 0.684 and 0.640, respectively. The minor alleles are *8,782T* and *28,144C*. Examination of seven SARS-CoV-2 strains collected in December 2019 revealed that they all carry these two major alleles, suggesting that they are ancestral alleles. On the evolutionary tree, the most recent common ancestor (MRCA) of SARS-CoV-2 is located at the root of the tree and found to harbor all of these ancestral alleles. The sequence between nucleotide position 100 and 29,800 of MRCA was found to be identical to that of the reference genome sequence (GenBank accession number: NC_045512)[3]. The finding is consistent with that of a previous study.[6]

### Construction of the evolutionary tree based on distributed alignments

To build the evolutionary tree, the sequence corresponding to the reference sequence between nucleotides 100 and 29,800 of each genome was used. Initially, the tree was built using the software FastTree[7] and a slightly revised version of RAxML[8]. To accommodate the entire length of each SARS-CoV-2 genome, the minimum branch length was changed from $10^{-5}$ to $10^{-10}$ in RAxML. However, these two methods were later found to be unsatisfactory because both FastTree and RAxML cannot analyze distributed alignments and sub-genomic regions. Furthermore, to use FastTree and RAxML, a unified multiple sequence alignment must be done for daily updates. This is beyond the capability of our computing facility. FastTree and RAxML also cannot distinguish missing bases from indels because both appear as “-” in the alignments. As gaps are ignored by these two methods and indels provide valuable information for construction of phylogenetic tree of closely related SARS-CoV-2 strains, new approaches are needed to accomplish the task. To simplify CGB implementation, the Neighbor-Joining method[9] was used.

When calculating genetic distances, five different features are considered. First, missing bases at 5’ and 3’ ends (presented as gaps in alignments) are ignored. Second, insertions and deletions are taken into consideration. Third, IUPAC (International Union of Pure and Applied Chemistry) ambiguous nucleotide characters (e.g., Y and R) are supported. As disambiguating nucleotides will generate a huge number of artificial sequences, genetic distances would be overestimated if all possible sequences are compared.

To solve this problem, the following strategy was used to treat ambiguous bases. For comparison of the sequence ACGRCG with the reference sequence ACGACG, ACGRCG is converted to ACGACG and ACGGCG. The resulting 2 new sequences are defined as one sequence set. Because this sequence set has the sequence ACGACG that is the same as that of the reference sequence, the strain with the sequence ACGRCG is considered as the same type as the strain with the reference sequence ACGACG. For comparison of the sequence ACGRCG with the sequence ACGYCG, ACGRCG is converted to ACGACG and ACGGCG, and ACGYCG is converted to ACGCCG and ACGTCG. Therefore, two sequence sets are generated. Because the four sequences in these two sequence sets are different, the strain with the sequence ACGRCG and the one with the sequence ACGYCG are considered as two different types. For comparison of the sequence ACGRCG with the sequence ACGHCG, ACGRCG is converted to ACGACG and ACGGCG, and ACGHCG is converted to ACGACG, ACGCCG and ACGTCG. As the resulting two sequence sets share the same sequence ACGACG, the strain with the sequence ACGRCG and the one with the sequence ACGHCG are considered as the same type.

Forth, the sequences of two genomes for comparison are placed in different alignments, and the sequence of the reference genome is used as the coordinate for nucleotide positions. Fifth, the genetic distance between outgroups and a SARS-CoV-2 strain is determined after adding two components: the average genetic distance between outgroups and the most recent common ancestor (MRCA), and the genetic distance between MRCA and the strain.

### Imputation of ambiguous and missing nucleotides

An ambiguous or missing base can be imputed (Figure S3) if the strain with the ambiguous base shares the same phylogeny with neighboring lineages[10]. For this imputation, the allele frequency and the definition of IUPAC ambiguous nucleotide characters are considered, and only the lineages with collection dates $\pm 30$ days apart are compared.

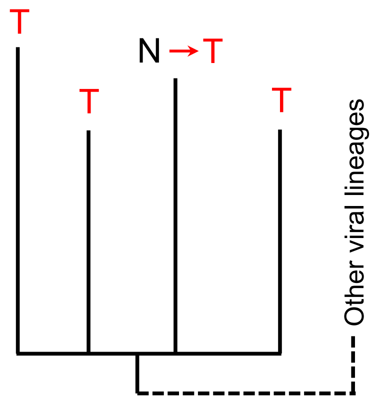

**Figure S3. Imputation of ambiguous nucleotides of a lineage using the information of its siblings.**

### Parsimony inference of mutations for strains in each branch

After ambiguous and missing nucleotides are replaced with inferred nucleotides, mutations in strains of each branch are recapitulated according to the principle of parsimony[11, 12]. Although the analysis is performed site by site, large deletions spanning over a number of regions are merged as a single large deletion, and a long insertion is considered as a united element. Thus it is easy to trace recurrent deletions[13] whenever necessary.

### Maximum-likelihood phylodynamic analysis

A highly effective maximum-likelihood method (TreeTime) is used to determine the dates of internal nodes[14] as it allows fast inference by "the post- and pre-order traversals" with tabulated key values for back tracing. This algorithm was implemented in CGB with very minor revisions. The genome-wide mutation rate is also timely updated to calculate the likelihood.

As recommended by TreeTime[14], all length zero branches are pruned, and branch length corresponds to number of mutations on the branch. To improve computation efficiency, CGB first categories branches with length zero according to its context (Figure S4). In some cases, branches with length zero are not pruned (Figure S4A, E) in order to make length zero offspring as a clade and to reduce the number of multifurcated nodes.

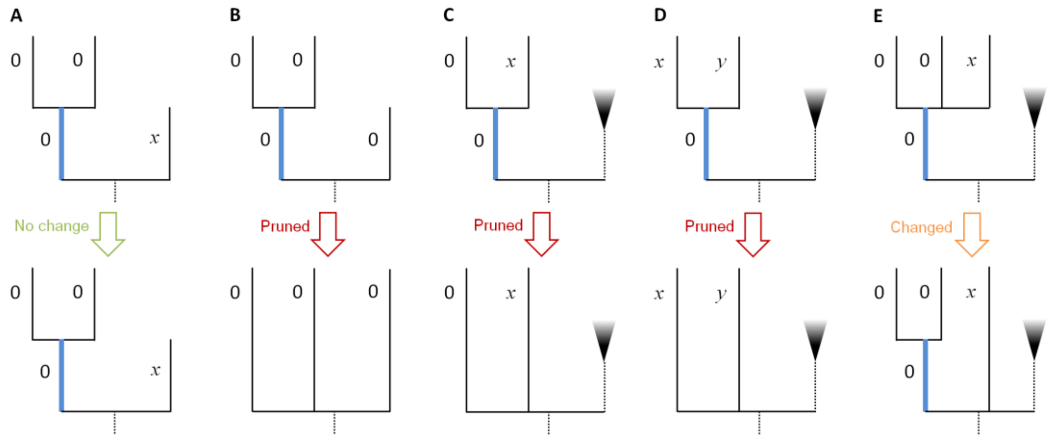

**Figure S4. Five categories of length zero branches (highlighted in blue).**

A) All offspring of the branch have length zero, and the sister branch of the branch has length non-zero *x*. In this case, the two offspring of length zero are in the same clade.

B) The sister branch has length zero, and the three nodes are clustered to form a multifurcated clade.

C) If one offspring of the branch has length zero, the branch is pruned.

D) If all offspring of the branch have length non-zero *x* or *y*, the branch is pruned.

E) If two or more offspring of the branch have length zero, the branch is kept and the non-zero branch is removed.

Many internal nodes are multi-furcated instead of bi-furcated because the viral strains are very similar to each other. The multi-furcated nodes are known as polytomies. To reduce the number of branches of a polytomic node, CGB sorts the branches according to the potential gain of likelihood if branches are shortened and determines whether a longer or shorter branch length would increase the likelihood of tree. The branches are bi-partitioned to form a new clade (Figure S5), and the two sets of branches are determined by maximizing the gain of likelihood. The bi-partition always starts from the root to the tips, and this process is repeated at least four times.

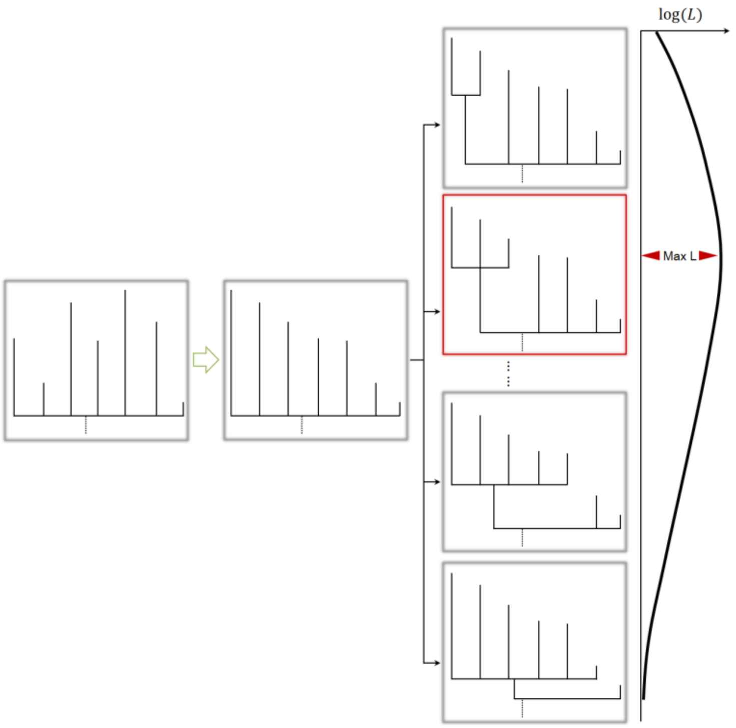

**Figure S5. Bi-partition of a polytomic node.** CGB first sorts the branches according to the potential gain of likelihood. If $k$ branches are linked to the node, there are $k-2$ different ways to bi-partition the node. The two sets of branches are determined by maximizing the gain of likelihood.

### Maximum-likelihood analysis based on the existing mutation-annotated tree

Branch and bound for maximum parsimony[15, 16] is implemented with a speed-up revision. New genomic sequences of SARS-CoV-2 strains are first aligned with the reference genome (Figure 1). The resulting alignment and previous results are then analyzed together, and the evolutionary tree is rebuilt using previous result file that contains the existing tree and mutation information. A new strain is then added to the mutation-annotated tree as a dated leaf, and new mutations are labeled and analyzed according to the principle of parsimony. CGB adds the earliest strain first to the tree. After adding all new genomic sequences, the mutation rate of SARS-CoV-2 is calculated, and the date of each internal node is determined as described above. This maximum-likelihood (ML) analysis was performed with a slightly revised version of TreeTime[14].

The speed-up-revised branch and bound provides a balance between efficiency and accuracy. However, it may not be globally optimized. To solve this problem, a sub-tree optimization is performed. As many internal branches have five or more mutations, the large evolutionary tree was divided into small subtrees. Because sub-tree optimization is much faster than rebuilding the whole tree, it is frequently performed as needed.

### Tree visualization with CGB

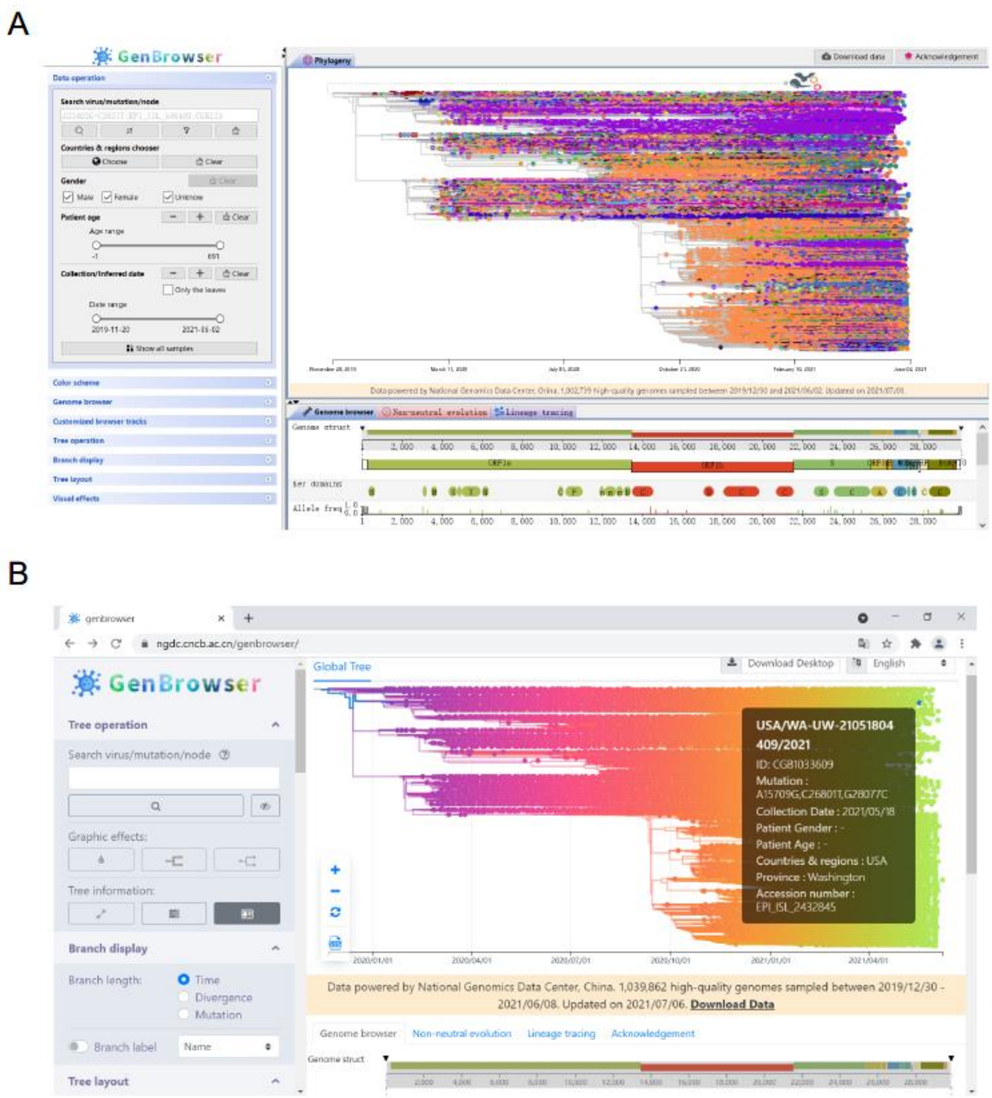

**Figure S6. Tree visualization with the CGB.**

A) Tree visualization in desktop version of CGB.

B) Tree visualization in web-based CGB. Nine language versions (Chinese, English, German, Japanese, French, Italian, Portuguese, Russian, and Spanish) are available.

**Table S1. Computation time for the rendering process (*n*=1,002,739) in two operation systems.**

| Operating System | Replicate | Calculation (ms) | First painting (ms) | Next paintings (ms) |
|---|---|---|---|---|
| MacOS Catalina 10.15.7 | 1 | 292 | 696 | 201 |
| 2.6 GHz four core Intel Core i7 | 2 | 235 | 709 | 200 |
| 16 GB 1600 MHz DDR3 | 3 | 293 | 762 | 203 |
| NVIDIA GeForce GT 750M 2 GB/Intel Iris Pro 1536 MB | 4 | 236 | 691 | 201 |
| Windows 10 Home version | 1 | 290 | 386 | 238 |
| 8GB RAM | 2 | 235 | 323 | 206 |
| Intel(R) Core(TM) i5-8250U | 3 | 298 | 376 | 197 |
| CPU @ 1.60GHz 1.80 GHz Intel(R) UHD Graphics 620 | 4 | 297 | 364 | 250 |

### Coordinated annotation tracks

CGB uses six tracks to show genome structure and key domains, allele frequencies, sequence similarity, multi-genome alignment and primer sets for detection of SARS-CoV-2 (Figure S7). These tracks are coordinated according to nucleotide positions of the SARS-CoV-2 reference genome.

The first track shows the structure of a SARS-CoV-2 genome. By dragging or right clicking the mouse, a genomic region can be zoomed in. The second track shows 25 known key domains. By right clicking on a domain box, amino acid sequence of the domain can be copied, and the related information page on the Pfam website (http://pfam.xfam.org) can be opened.

The third track shows the frequencies of derived alleles or variants (Figure S7). Since the web version is designed for the general public and quick view of global samples, users can update manually the frequency of an allele in the chosen clade. When hovering mouse on the frequency column of an allele, its allele frequency trajectory (Figure S8) will pop up. This allele frequency trajectory is calculated by a sliding window of five days in size. The person who first discovered the allele is indicated below allele frequency trajectory.

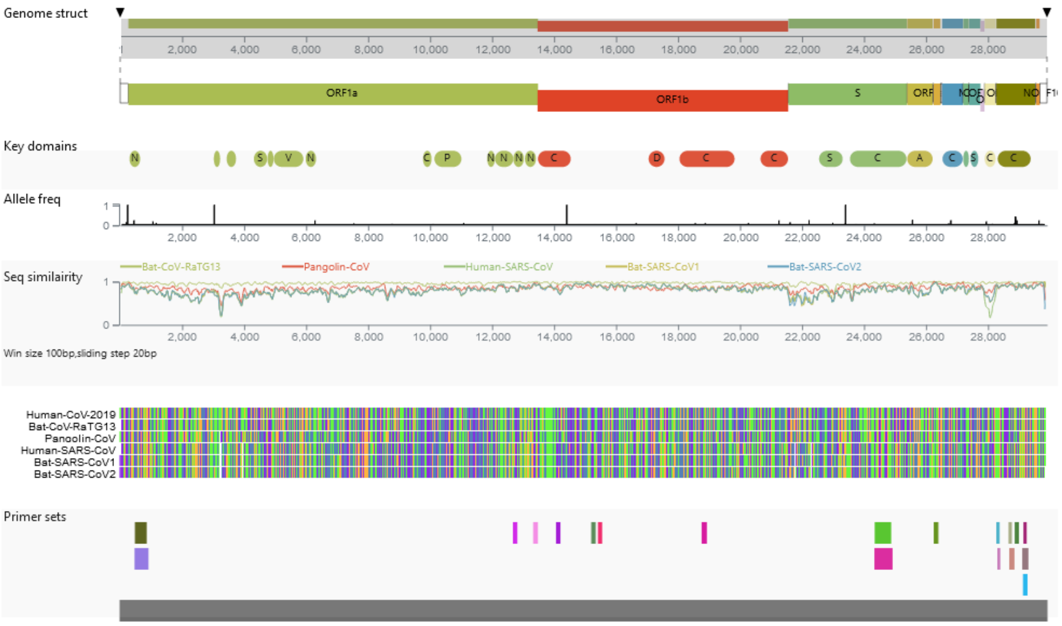

**Figure S7. Six tracks shown by the Coronavirus GenBrowser.**

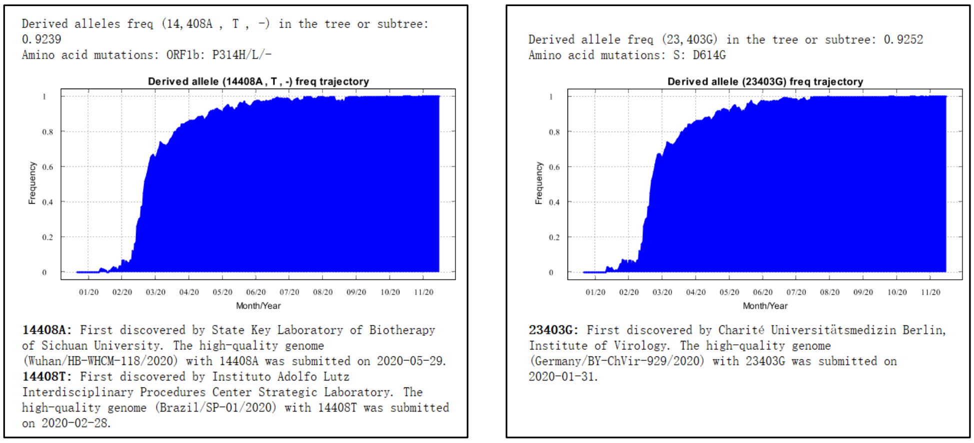

**Figure S8. Visualization of allele frequency trajectory with CGB.**

The fourth track shows sequence similarity between SARS-CoV-2 reference genome (NC_045512.2)[3] and the genomes of five other coronaviruses, including bat-CoV-RaTG13 (MN996532.1)[4], pangolin-CoV (MT040334.1)[5], human-SARS-CoV (AY278488.2)[17], bat-SARS-CoV1 (KY417146.1)[18], and bat-SARS-CoV2 (MK211376.1)[19]. Sequence similarity is determined using a sliding window (window size 100 bases and sliding step 20 bases). In the standalone version of CGB, these parameters can be adjusted to re-calculate the degree of sequence similarity.

The fifth track shows alignments of six coronaviruses performed with the software MAFFT[2]. Nucleotide sequences of five coronaviruses are coordinated according to nucleotide positions of the SARS-CoV-2 reference genome. Inserted sequences, if any, in the genomes of the five non-SARS-CoV-2 coronaviruses can be viewed with the standalone version of CGB (Figure S9).

The sixth track presents primer sets that can be used to detect various SARS-CoV-2 genes or strains. Various regions of the genome that can be amplified are indicated. Combined with allele frequency information, the efficiency of nucleic acid testing can be verified (Figure S10). Since viral strains can be filtered according to collection dates and locations, their allele frequency can be easily determined.

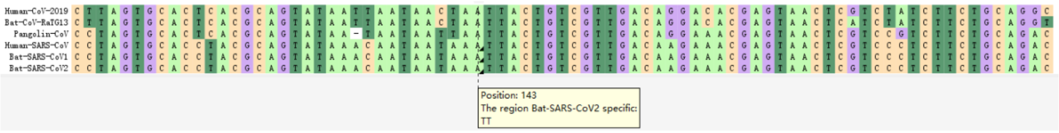

**Figure S9. Multiple-genome alignment.** Inserted sequences in the five non-SARS-CoV-2 genomes are marked with black triangles. This alignment can be downloaded from https://bigd.big.ac.cn/ncov/apis/.

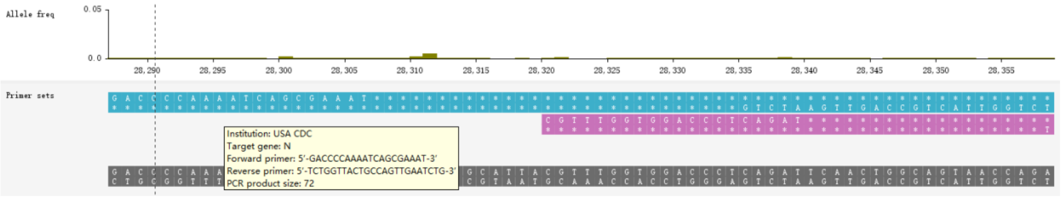

**Figure S10. Combined view of two tracks of allele frequency and primer set for detection of various SARS-CoV-2 strains.** The nucleotide sequences of two primers are shown, and their amplified region is marked in pink.

### Data searching, filtering, and visualization of a single clade on the huge tree

To view a lineage on the huge evolutionary tree, several different data searching and filtering methods can be used. A clade can be viewed in a new tab, and its sub-clade can be viewed in another new tab. A clade can also be collapsed or un-collapsed. Moreover, chosen lineages can be made visible, and un-chosen ones can be hidden. After right clicking a branch, a menu will pop up to help navigate through the huge tree. A lineage can also be viewed by deep zoom-in using the desktop standalone version of CGB (Figure 2A). However, the deep zoom-in function is not implemented in the web-based CGB because it is a simplified version and is designed mainly for educational purpose.

### CGB binary nomenclature for each internal node or branch

To name each internal node or branch, the CGB binary nomenclature system was developed following the MRCA concept as follows. Each node of a viral strain is first assigned a permanent unique positive integer (e.g., 1 – 9) in the order of discovery (Figure S11). Assuming that an internal node has two sub-nodes that are named CGB1 and CGB2, this internal node is named CGB1.2. For an internal node with more than two sub-nodes, e.g., CGB7, CGB9, and CGB6, it is named with the two smallest CGB numbers, given the condition that the internal node is the MRCA of the two sub-nodes, separated by a dot; thus, this internal node is designated as CGB6.7.

This naming process is very fast, and all nodes of the huge evolutionary tree can be named in seconds. Each node can be easily searched and viewed by CGB. When a new sequence is added to the tree as a sub-node, its CGB number would be greater than all the pre-existing CGB numbers and thus will not change the previously assigned CGB number of the internal node, which the new sequence belongs.

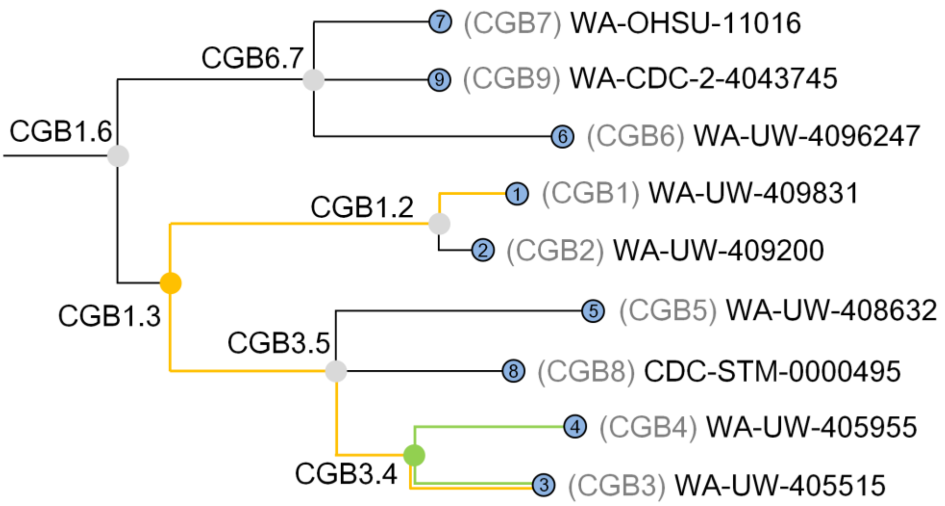

**Figure S11. Illustration of CGB binary nomenclature.** The evolutionary tree is shown with nine strains indexed CGB1 – CGB9. Their pseudo-isolate names are shown. The green internal node with two sub-nodes named CGB3 and CBG4 is designated as CGB3.4 since the MRCA of CGB3 and CGB4 is the green node. For an internal node with more than two sub-nodes, it is named with the two smallest CGB numbers, given the condition that the internal node is the MRCA of the two sub-nodes, separated by a dot. Therefore, an orange internal node is named CGB1.3 because it contains CGB1, CGB2, CGB5, CGB8, CGB4, and CGB3 with CGB1 and CGB3 being the smallest CGB numbers, on the condition just described.

### Estimation of mutation rate

The 95% confidence interval of the estimated mutation rate is obtained via Monte-Carlo simulations. Given the estimated mutation rate, mutations are randomly generated along the evolutionary tree[20], and mutation rate is estimated by regression analysis. Then the empirical distribution of estimated mutation rate is obtained from 1,000 simulated data set.

**Table S2. Mutation rate of various SARS-CoV-2 genes.**

| SARS-CoV-2 Gene | Mutation rate (per nucleotide per year) |
|---|---|
| ORF1a | $5.9704 \times 10^{-4}$ |
| ORF1b | $4.774 \times 10^{-4}$ |
| S | $2.4343 \times 10^{-3}$ |
| ORF3a | $3.2810 \times 10^{-5}$ |
| E | $5.4687 \times 10^{-5}$ |
| M | $1.0794 \times 10^{-3}$ |
| ORF6 | $1.6243 \times 10^{-4}$ |
| ORF7a | $6.7185 \times 10^{-4}$ |
| ORF7b | $4.5438 \times 10^{-4}$ |
| ORF8 | $9.4007 \times 10^{-3}$ |
| N | $4.9052 \times 10^{-3}$ |
| ORF10 | $1.0794 \times 10^{-3}$ |
| noncoding | $3.5758 \times 10^{-3}$ |

### Mutations affected by recombination

To determine the effect of recombination on evolution, it is necessary to understand the history of recombination which is usually represented by the ancestral recombination graph (ARG)[21-23]. Because it is impossible to construct an ARG for the huge collection of SARS-CoV-2 variants, a new method needs to be developed. According to the finite sites model, which is commonly used to study fast evolving organisms[24], recombination and recurrent mutation can generate similar genomic variants (Figure S8). As recombination creates a hybrid genomic structure[25], it can be distinguished from a recurrent mutation (Figure S12), which affects only the mutated site. In contrast, a recombination event affects a large part of genome (Figure S12A).

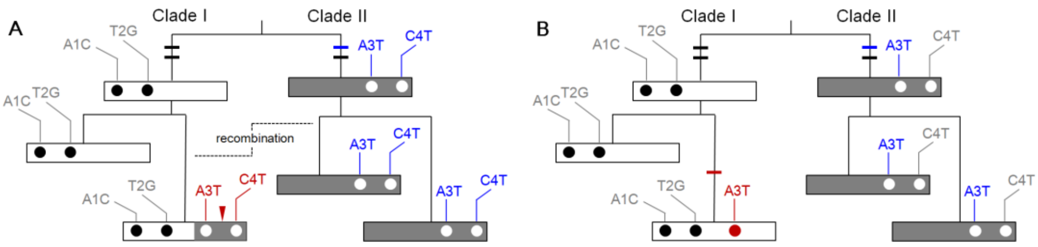

**Figure S12. Generation of similar genomic variants by recombination and recurrent mutation.**

A) Recombination creates a hybrid genomic structure. The region affected by recombination is indicated with a red arrowhead. Each notch of the branches represents a mutation. Open and dark gray square strips represent sequences in two lineages. Solid and empty circles denote mutations. In clade I, mutations A3T and C4T are observed due to recombination. These two mutations are considered recurrent if the recombination is ignored because they are also present in clade II.

B) A recurrent mutation A3T, marked in red, occurs in clade I. The same mutation (marked in blue) also occurs in clade II.

To identify mutations due to recombination, a flagging procedure is performed in four steps. First, multiple mutations that occur at the same genomic position, all mutations are labeled with a recombination flag. Second, mutations are categorized according to their types. Different mutations are considered as the same type if their ancestral and derived alleles are the same. Third, for each category, the recombination flag of the most prevalent mutation is removed because this mutation is unlikely caused by recombination. The prevalence of a certain mutation corresponds to the number of its descendants[26] Back mutations are not considered. Forth, if two recombination-flagged mutations are less than 20 kb apart, their recombination flags are maintained.

### Identification of mutation cold spots

To find mutation cold spots, the mutation density of a genome is denoted as $\beta$ (mutations per base), and the observed number of mutations within a 10-base window is denoted as $\xi_{obs}$. Under the assumption of homogeneous mutation distribution, the expected number of mutations within the window is $10\beta$. The significant level of mutation cold spots is determined by Poisson probability[27, 28]: $P(x \leq \xi_{obs}) = \sum_{x \leq \xi_{obs}} e^{-10\beta} (10\beta)^x / x!$. It is a one-tailed test. Since a deletion may include multiple bases, the number of deleted bases, instead of the number of deletions, is used to determine the Poisson probability. If insertions are present, the window is ignored. Finally, overlapped windows are merged to form a mutation cold spot.

Among the identified cold spots, those conserved in SARS-CoV-2 but not conserved in other coronaviruses were identified. The sequence similarity (window size 100 bases and sliding step 20 bases) was calculated between SARS-CoV-2 (NC_045512)[3] and each of five coronaviruses (Bat-CoV-RaTG13[4], Pangolin-CoV[5], Human-SARS-CoV[17], Bat-SARS-CoV1[18], Bat SARS-CoV2[19]). If the average sequence similarity of a region is smaller than 70%, the region is treated as non-conserved in the coronaviruses.

### Detection of on-going selection of SARS-CoV-2

To detect on-going positive selection, allele frequency trajectory with an S-shaped curve is examined as described previously[29-32]. For this determination, the selection coefficient is denoted as $s$. The initial frequency of the derived allele $a$ is denoted as $q_0$, and that of the wide-type allele $A$ is $p_0 = 1 - q_0$. The frequency of the wild-type allele $A$ at a specific day (time $t$) is $p_t$, and that of the derived allele $a$ is $q_t$.

The following equation was used to calculate the coefficient of on-going positive selection[29] (Table S3):

$$\frac{q_t}{p_t} = (1+s)\frac{q_{t-1}}{p_{t-1}} = \cdots\cdots = (1+s)^t\frac{q_0}{p_0}. \quad (1)$$

Then

$$log\left(\frac{q_t}{p_t}\right) = log\left(\frac{q_0}{p_0}\right) + t log(1+s). \quad (2)$$

Since $t$ is known, $log(1+s)$ can be estimated by linear regression.

**Table S3. Frequency of wild type and derived alleles after selection.**

| Haplotype | $A$ (wild type) | $a$ (derived allele) |
|---|---|---|
| Fitness | 1 | $1+s$ |
| Frequency at the $(t-1)$-th day | $p_{t-1}$ | $q_{t-1}$ |
| Frequency at the $t$-th day | $p_t = \frac{p_i}{p_i + (1+s)q_i}$ | $q_t = \frac{(1+s)q_i}{p_i + (1+s)q_i}$ |

As shown in Figure S13, the best time window to control the transmission of a strain with an advantageous mutation is shadowed. When $s > 0$, the frequency of a derived allele increases over time[29]. During Stages I and III, the speed of increase in the frequency of advantageous allele is slow, indicating low selection efficiency. During Stage II, the speed of increase in the frequency of advantageous allele is fast, and the efficiency of selection is high. When the frequency is 50%, the efficiency of selection reaches maximum. Therefore, the best time window to control the transmission of strains with an advantageous mutation is that of Stage I, especially when its frequency is still below 10%.

The analysis framework for detecting strains with putative advantageous mutations during their early stage of spreading is summarized in Figure S14. A neutral mutation may be linked to an advantageous mutation and spread over the entire population[21, 22, 33]. To reduce the impact of hitchhiking by neutral mutation, only non-synonymous mutations were analyzed. For this analysis, the initial (start) frequency must be < 0.1, and the end frequency must be > 0.05. Only the mutation frequency trajectory during the selective phase was used for calculation as this is the period when an advantageous mutation causes on-going selection. Linear regression analysis was performed to detect advantageous mutation. According to the equation described above, a mutation was considered advantageous when $s > 0$, $p < 0.01$, and $R^2 > 0.5$.

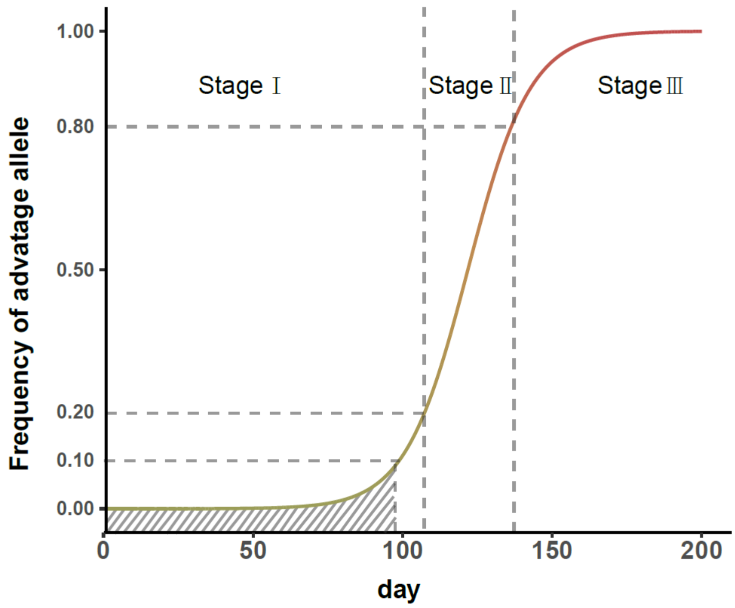

**Figure S13. S-shaped frequency trajectory of advantageous mutations.** $s = 0.1$ and $q_0 = 0.0001$.

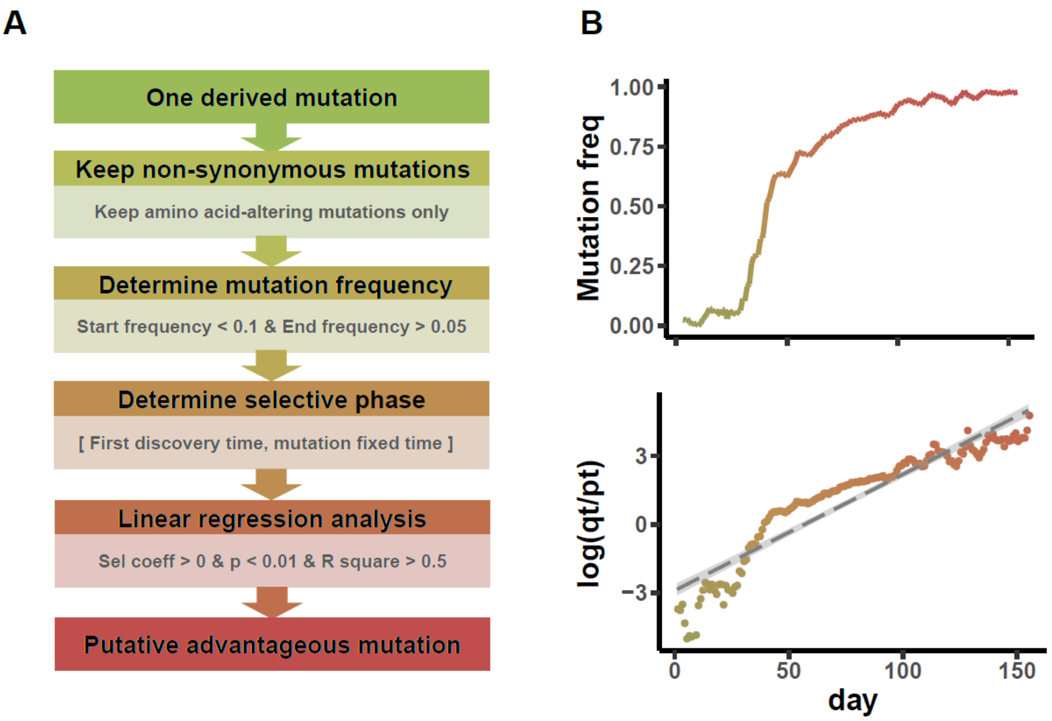

**Figure S14. Detection of on-going selection of SARS-CoV-2.**
**A)** Flow chart for detection of putative advantageous variants.
**B)** Frequency trajectory for A23403G (S: D614G) and linear regression analysis. The *x*-axis displays number of days since the first appearance of a derived allele in global virus population. $q_t$ is the frequency of the derived allele (*23403G*), and $p_t$ is the frequency of the ancestral allele (*23403A*) at time *t*.

**Table S4. Putative advantageous mutations in the spike protein.**¶

| Position | Nucl. mut. | AA mut. | Start time | Start freq | End/Last time | End/Last freq | Sel Coeff | P-value | R-square |
|---|---|---|---|---|---|---|---|---|---|
| 21765* | TACATG21765- | HV69- | 2020/3/26 | 0.0003 | 2021/5/28 | 0.4439 | 0.0265 | <1.0E-10 | 0.9291 |
| 21991* | TTA21991- | Y144- | 2020/2/16 | 0.0068 | 2021/5/28 | 0.432 | 0.0238 | <1.0E-10 | 0.7969 |
| 22917 | T22917G | L452R | 2020/3/15 | 0.0002 | 2021/5/28 | 0.5408 | 0.0175 | <1.0E-10 | 0.8314 |
| 22995 | C22995A | T478K | 2020/4/26 | 0.0004 | 2021/5/28 | 0.5357 | 0.02 | <1.0E-10 | 0.7645 |
| 23063* | A23063T | N501Y | 2020/3/28 | 0.0002 | 2021/5/28 | 0.449 | 0.0305 | <1.0E-10 | 0.8757 |
| 23271* | C23271A | A570D | 2020/4/25 | 0.0004 | 2021/5/28 | 0.4388 | 0.0362 | <1.0E-10 | 0.7822 |
| 23403 | A23403G | D614G | 2020/1/17 | 0.0263 | 2020/7/21 | 0.9913 | 0.046 | <1.0E-10 | 0.8667 |
| 23604* | C23604A | P681H | 2020/3/25 | 0.001 | 2021/5/28 | 0.4405 | 0.0271 | <1.0E-10 | 0.9173 |
| 23604 | C23604G | P681R | 2020/6/26 | 0.003 | 2021/5/28 | 0.5374 | 0.0183 | <1.0E-10 | 0.697 |
| 23709* | C23709T | T716I | 2020/3/25 | 0.0002 | 2021/5/28 | 0.4388 | 0.0282 | <1.0E-10 | 0.8519 |
| 24506* | T24506G | S982A | 2020/9/18 | 0.0004 | 2021/5/28 | 0.4388 | 0.0377 | <1.0E-10 | 0.7789 |
| 24914* | G24914C | D1118H | 2020/3/31 | 0.0002 | 2021/5/28 | 0.4388 | 0.0355 | <1.0E-10 | 0.7964 |

¶The analysis was performed on global samples ($n = 1{,}002{,}739$).
*Mutations found on the clade CGB84017.91425 (B.1.1.7).

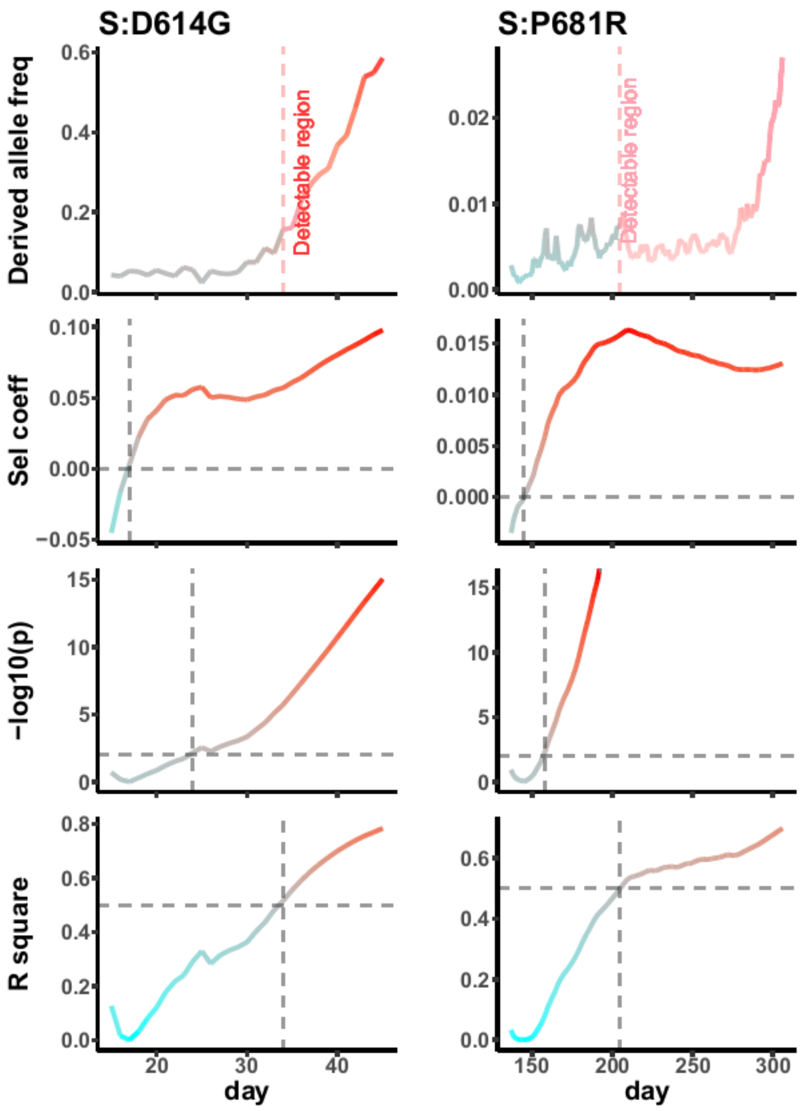

**Figure S15. Putative advantageous variants of SARS-CoV-2.** The x-axis displays number of days since the first appearance of derived allele in the global viral population. Predicted adaptation is marked in pink. Dashed gray crossings denote top right corners with a positive selection coefficient, $p<0.01$, and R^2>50%.

### References

1. Zhao W-M, Song S-H, Chen M-L et al. The 2019 novel coronavirus resource, Hereditas (Beijing) 2020;42:212-221.
2. Rozewicki J, Li S, Amada KM et al. MAFFT-DASH: integrated protein sequence and structural alignment, Nucleic Acids Res 2019;47:W5-W10.
3. Wu F, Zhao S, Yu B et al. A new coronavirus associated with human respiratory disease in China, Nature 2020;579:265-269.
4. Zhou P, Yang X-L, Wang X-G et al. A pneumonia outbreak associated with a new coronavirus of probable bat origin, Nature 2020;579:270-273.
5. Lam TT-Y, Jia N, Zhang Y-W et al. Identifying SARS-CoV-2-related coronaviruses in Malayan pangolins, Nature 2020;583:282-285.
6. Bedford T, Greninger AL, Roychoudhury P et al. Cryptic transmission of SARS-CoV-2 in Washington state, Science 2020;370:571-575.
7. Price MN, Dehal PS, Arkin AP. FastTree 2-Approximately maximum-likelihood trees for large alignments, Plos One 2010;5:e9490.
8. Kozlov AM, Darriba D, Flouri T et al. RAxML-NG: a fast, scalable and user-friendly tool for maximum likelihood phylogenetic inference, Bioinformatics 2019;35:4453-4455.
9. Saitou N, Nei M. The neighbor-joining method: a new method for reconstructing phylogenetic trees, Mol Biol Evol 1987;4:406-425.
10. Li H, Zhang YW, Zhang YP et al. Neutrality tests using DNA polymorphism from multiple samples, Genetics 2003;163:1147-1151.
11. Sankoff D. Minimal mutation trees of sequences., SIAM J Appl Math 1975;28:35-42.
12. Hartigan JA. Minimum mutation fits to a given tree, Biometrics 1973;29:53-65.
13. McCarthy KR, Rennick LJ, Nambulli S et al. Recurrent deletions in the SARS-CoV-2 spike glycoprotein drive antibody escape, Science 2021:eabf6950.
14. Sagulenko P, Puller V, Neher RA. TreeTime: Maximum-likelihood phylodynamic analysis, Virus Evol 2018;4:vex042.
15. White WTJ, Holland BR. Faster exact maximum parsimony search with XMP, Bioinformatics 2011;27:1359-1367.
16. Hendy MD, Penny D. Branch and bound algorithms to determine minimal evolutionary trees, Math Biosci 1982;59:277-290.
17. Qin E, Zhu QY, Yu M et al. A complete sequence and comparative analysis of a SARS-associated virus (Isolate BJ01), Chin Sci Bull 2003;48:941-948.
18. Hu B, Zeng L-P, Yang X-L et al. Discovery of a rich gene pool of bat SARS-related coronaviruses provides new insights into the origin of SARS coronavirus, PLoS Pathog 2017;13:e1006698.

19. Han YL, Du J, Su HX et al. Identification of diverse bat alphacoronaviruses and betacoronaviruses in China provides new insights into the evolution and origin of coronavirus-related diseases, Front Microbiol 2019;10:1900.
20. Li H, Stephan W. Inferring the demographic history and rate of adaptive substitution in *Drosophila*., PLoS Genet. 2006;2:e166.
21. Kim Y, Stephan W. Detecting a local signature of genetic hitchhiking along a recombining chromosome., Genetics 2002;160:765-777.
22. Li H, Stephan W. Maximum likelihood methods for detecting recent positive selection and localizing the selected site in the genome., Genetics 2005;171:377-384.
23. Bouckaert R, Vaughan TG, Barido-Sottani J et al. BEAST 2.5: An advanced software platform for Bayesian evolutionary analysis, PLoS Comput Biol 2019;15.
24. Gao F, Ming C, Hu WJ et al. New software for the fast estimation of population recombination rates (FastEPRR) in the genomic era, G3 2016;6:1563-1571.
25. Lam HM, Ratmann O, Boni MF. Improved algorithmic complexity for the 3SEQ recombination detection algorithm, Mol Biol Evol 2018;35:247-251.
26. Fu Y-X. Statistical properties of segregating sites., Theor Popul Biol 1995;48:172-197.
27. Ohta T, Kimura M. On the constancy of the evolutionary rate in cistrons, J Mol Evol 1971;1:18-25.
28. Wang Y, Dai G, Gu Z et al. Accelerated evolution of an *Lhx2* enhancer shapes mammalian social hierarchies, Cell Res 2020;30:408-420.
29. Hartl DL, Clark AG. Principles of Population Genetics. Sunderland, Massachusetts: Sinauer Associates, Inc., 1988.
30. Li J, Schneider KA, Li H. The hitchhiking effect of a strongly selected substitution in male germline on neutral polymorphism in a monogamy population, Plos One 2013;8:e71497.
31. Stephan W, Wiehe THE, Lenz MW. The effect of strongly selected substitutions on neutral polymorphism: analytical results based on diffusion theory., Theor. Popul. Biol. 1992;41:237-254.
32. Schraiber JG, Evans SN, Slatkin M. Bayesian inference of natural selection from allele frequency time series, Genetics 2016;203:493-511.
33. Kaplan NL, Hudson RR, Langley CH. The "hitchhiking effect" revisited., Genetics 1989;123:887-899.
